# A Single Anodal Transcranial Direct Current Stimulation Session to Enhance Sport-Specific Performance in Trained Individuals? A Systematic Review and Meta-Analysis

**DOI:** 10.1101/2022.06.23.22276798

**Authors:** Tom Maudrich, Patrick Ragert, Stephane Perrey, Rouven Kenville

**Author notes:** **Correspondence:** Tom Maudrich. **Author Contributions** PR provided the idea of the systematic review and meta-analysis. TM & RK independently performed the literature search and meta-analysis. TM, PR, SP & RK wrote the manuscript. All authors interpreted the data, contributed to the manuscript, reviewed, approved the content of the final version, and agree to be accountable for all aspects of the work. All persons designated as authors qualify for authorship, and all those who qualify for authorship are listed.

## Abstract

**Background:** Transcranial direct current stimulation (tDCS) has emerged as a promising and feasible method to improve motor performance in healthy and clinical populations. However, the potential of tDCS to enhance sport-specific motor performance in athletes remains elusive.

**Objective:** We aimed at analyzing the acute effects of a single anodal tDCS session on sport-specific motor performance changes in athletes compared to sham.

**Methods:** A systematic review and meta-analysis was conducted in the electronic databases PubMed, Web of Science, and SPORTDiscus with publication dates through April 2022. The meta-analysis was performed using an inverse variance method and a random-effects model. Additionally, two subgroup analyses were conducted (1) depending on the stimulated brain areas (i.e., primary motor cortex (M1), temporal cortex (TC), prefrontal cortex (PFC), cerebellum (CB)), and (2) studies clustered in subgroups according to different sports performance domains (endurance, strength, and visuomotor skill).

**Results:** A total number of 18 studies enrolling a sample size of 245 athletes were deemed eligible for inclusion. Across all included studies, a significant moderate standardized mean difference (SMD) favoring anodal tDCS to enhance sport-specific motor performance could be observed (SMD = 0.27, 95%CI [0.10, 0.44], p = 0.002). Subgroup analysis depending on cortical target areas of tDCS indicated non-significant moderate to high SMD in favor of anodal tDCS compared to sham for M1 (SMD = 0.24, 95%CI [-0.01, 0.49], p = 0.06), TC (SMD = 0.40, 95%CI [-0.10, 0.89], p = 0.12), PFC (SMD = 0.23, 95%CI [-0.04, 0.50], p = 0.09) and CB (SMD = 0.89, 95%CI [-0.15, 1.94], p = 0.09). Performance domain subgroup analysis revealed non-significant moderate SMD favoring anodal tDCS compared to sham: endurance domain (SMD = 0.23, 95%CI [-0.01, 0.47], p = 0.06), strength domain (SMD = 0.44, 95%CI [-0.14, 1.01], p = 0.14, Chi^2^ = 0.31) and visuomotor skill domain (SMD = 0.30, 95%CI [-0.03, 0.62], p = 0.07).

**Conclusion:** A single anodal tDCS session leads to performance enhancement in athletes in sport-specific motor tasks. Although no conclusions can be drawn regarding the modes of action as a function of performance domain or stimulation site, these results imply intriguing possibilities concerning sports performance enhancement. Furthermore, this study highlights the need to investigate tDCS applications under real-life conditions rather than in highly controlled laboratory settings to uncover the true potential of non-invasive brain stimulation as a performance enhancement not only in sports but also in the context of prevention or rehabilitation of neurological diseases.

## 1 Introduction

Top athletic performance arises from optimal integration of physical and mental capacities, both of which can be trained and improved through appropriate interventions. At the core of such interventions, especially in the physical domain, often lies the refinement of neural information processing, i.e., facilitation of sensory input, filtering of relevant stimuli, and streamlining motor responses [1]. In addition to traditional physical training, non-invasive brain stimulation (NIBS) has emerged as a potential performance-enhancing tool over the past decades [2]. Although a number of NIBS techniques have been developed to modulate cortical processing, transcranial direct current stimulation (tDCS) is one of the more promising and feasible strategies to enhance athletic performance [3]. This is due to safety issues, the simplicity of implementation, lack of interference during task execution, and comparably low costs [2]. tDCS involves the application of a subthreshold current to the brain, which, depending on the polarity (anodal or cathodal stimulation), leads to an increase (anodal) or decrease (cathodal) in the excitability of the cortical areas underneath the stimulation electrode [4]. tDCS has been successfully employed in healthy and clinical populations. Both cognitive and motor functions could be improved in healthy adults, and partially restored in patients suffering from Parkinson’s disease or stroke (for a detailed description of these findings, please see the following reviews [5-8]).

The fact that tDCS has been reported to be a potential performance-enhancing tool in several domains has focused attention on enhancing physical performance in sports (please see the following opinion articles for further reading [9, 10]). Particularly in the context of motor functions, there is a growing interest to evaluate the potential of tDCS in high-performance populations, i.e., athletes. This is evident, among other things, in the recent increase in reviews and meta-analyses evaluating the enhancement of motor skills by tDCS in healthy individuals (for further reading please refer to [11-13]). Initial positive findings led to the now-common term *Neurodoping* [9]. Interestingly, many of such findings related to increases in motor and cognitive functions in healthy, fit, but not athletically active individuals. Hence, evidence for tDCS-induced performance enhancement among athlete groups remains elusive. Currently, only a limited number of studies have evaluated tDCS-induced performance enhancements in athletes. Results are mixed, with some studies showing improvements in a variety of physical performance measures [14, 15] while others did not find significant changes [16] or even deterioration [17]. A critical aspect of studies on performance enhancement of athletes using brain stimulation is the choice of motor task. Many studies aimed to increase general conditional abilities, i.e., strength and endurance through tDCS [18, 19]. Others investigated the effects of tDCS on abstracted motor tasks, e.g., serial reaction time tasks or finger tapping tasks [20]. However, few studies have investigated whether or not tDCS is capable of improving sport-specific motor performance in athletes. Oftentimes, controlled, stationary motor tasks unrelated to the specific need in a sports discipline, were investigated in laboratory settings, e.g., visually cued reaction times in football and handball athletes [21]. To truly investigate tDCS effects on performance enhancement in athletes, ideally, motor tasks should be studied that have a high overlap with the sport of the investigated athletes. First, this increases the likelihood of performance improvement of an already high-level athlete since positive motor transfer, i.e., the degree to which motor performance in one task can be transferred to another task, is related to task-related experience [22-24]. On the other hand, such sport-specific tasks are more valuable because they relate to the athlete’s sport and therefore have higher relevance to the sport than tasks that test general motor skills.

As noted above, reviews and meta-analyses already exist that highlight the potential of tDCS to enhance performance in the motor domain [11-13]. However, none of these studies have focused on sport task specificity in athletes. Therefore, the present work aims to address this question, by conducting a systematic review and meta-analysis of studies that investigated acute effects of anodal tDCS on sport-specific performance changes in athletes. Here, the term sport-specific motor tasks denote such tasks that have a high overlap with motor tasks in the sport of the investigated athlete groups. Peak athletic performance is highly specific. In this sense, the investigation of sport-specific performance changes through tDCS seems essential to further approach the understanding of the potential of tDCS applications under real-life conditions compared to highly controlled laboratory settings. Such evidence might also have relevance in the context of neuromodulation in neurorehabilitation or prevention.

## 2 Materials and Methods

The systematic review and meta-analysis were conducted following the guidelines and recommendations contained in the PRISMA 2020 statement [25] and according to Cochrane guidelines [26].

### 2.1 Eligibility criteria

Studies were deemed eligible for analysis according to the PICOS inclusion criteria [27] if they contained the following factors:

- Population: healthy male or female adult athletes (participating regularly in organized sport for at least 2 years before the experiment), free of injury or neuronal disease
- Intervention: acute effects of a single anodal tDCS session on sport-specific motor performance
- Comparator: sham stimulation in a single or double-blind design
- Outcomes: performance in sport-specific motor tasks
- Study design: randomized-controlled trials (RCT) with crossover or parallel design Articles that did not meet the inclusion criteria were excluded from this systematic review and meta-analysis.

### 2.2 Information sources

A systematic literature search was performed by two independent researchers (TM, RK) in the electronic databases PubMed, Web of Science, and SPORTDiscus with publication year until April 2022. The reference lists of the included studies were also scanned to generate a broader scope of the search. Only studies published in the English language were reviewed and included in the systematic review and meta-analysis.

### 2.3 Search strategy

Searches were performed in PubMed (all fields), Web of Science (all fields), and SPORTDiscus (all fields) using the keywords “transcranial direct current stimulation” OR “tDCS” AND “athletes”.

### 2.4 Selection process

Records were screened and selected by two review authors (TM, RK) independently based on previously defined PICOS eligibility criteria (see flow diagram Figure 1). Disagreements were resolved by reaching a consensus or by involving a third person (PR).

**Figure 1.**
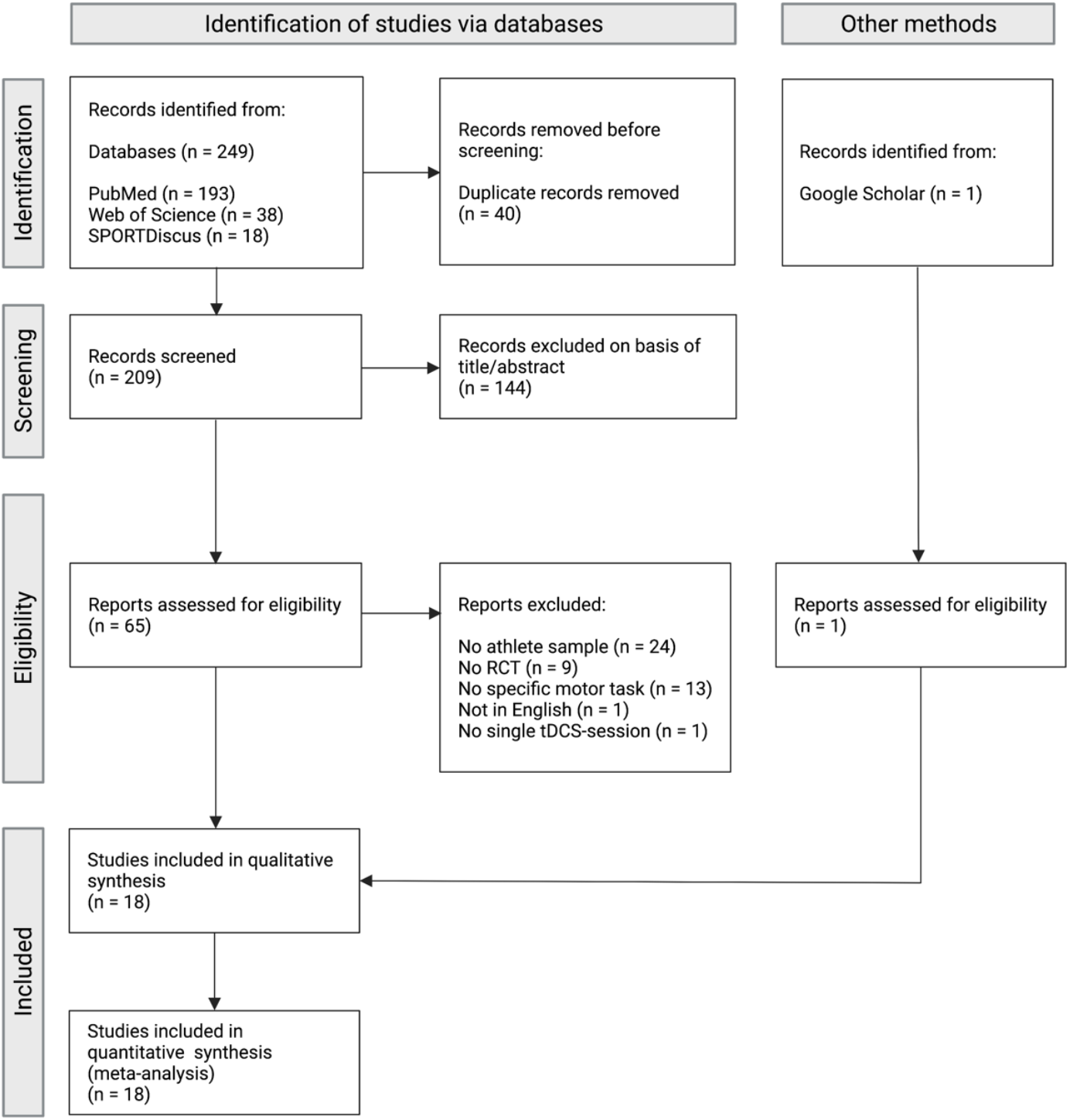
PRISMA flow chart diagram depicting the study selection process. Initially, 249 records were identified of which 18 studies were deemed eligible within the scope of the qualitative and quantitative synthesis.

### 2.5 Data extraction

Two review authors (TM, RK) independently extracted the following data items from the included studies:

1. Methods: study design (crossover/parallel RCT).

2. Participants: number, gender, sports discipline, training experience

3. tDCS application: tDCS electrode location, stimulation intensity, stimulation density, stimulation duration, motor task during/after stimulation, high-definition (HD)-tDCS or conventional tDCS.

4. Outcomes: sport-specific motor tasks.

5. Notes: funding for studies and notable conflicts of interest of authors.

Disagreements were resolved by reaching a consensus or by involving a third person (PR). If data were not reported within a manuscript, the authors of the original papers were contacted or values were extracted using Webplot Digitizer version 4.4 (https://apps.automeris.io/wpd/).

### Risk of bias assessment

Risk of bias assessment for randomized trials was performed by two review authors (TM, RK) independently according to the criteria contained in the Cochrane guidelines [28]: (1) random sequence generation (selection bias) (2) allocation concealment (selection bias), (3) blinding of participants and personnel (performance bias),(4) blinding of outcome assessment (detection bias), (5) incomplete outcome data (attrition bias), (6) selective reporting (reporting bias), and (7) “other bias.” For every included study, each of these items was classified as “low risk of bias” (“+”), “high risk of bias” (“-”), or “unclear risk of bias” (“?”). For this purpose, the software Review Manager 5.4.1 (Cochrane Collaboration, Oxford, UK) was used. Any disagreements in ratings of risk of bias were handled by a conversation between the two evaluators and consultation with a third person (PR).

### 2.6 Quantitative Analysis

The meta-analysis was performed using Review Manager 5.4.1 (Cochrane Collaboration, Oxford, UK). The intervention effects of tDCS on sport-specific motor task changes were calculated within each study using the standardized mean difference (SMD) of the continuous data at a 95% confidence interval (95%CI). Therefore, the mean difference and standard deviation in performance between anodal tDCS and sham were calculated in a sport-specific motor task closely related to competition in the sports discipline (e.g., cycling; 20 min time trial on a bicycle ergometer). In the case of multiple motor tasks performed within one study, the motor task that best represents competition performance in the respective athlete population was selected (for selected sport-specific tasks, please see Table 1). For tasks in which reaction times or time trials were assessed, mean outcome values were multiplied by -1 to ensure that all intervention effects pointed in the same direction (i.e., lower reaction time means better performance). Because the included studies used different sport-specific motor tasks in various populations of trained individuals, SMDs were weighted by the inverse variance method, and a random-effects model was used to account for statistical heterogeneity and to minimize the imprecision of the pooled effect estimate. Studies were clustered in subgroups depending on the brain area stimulated by tDCS (i.e., primary motor cortex (M1), temporal cortex (TC), prefrontal cortex (PFC), cerebellum (CB)). Furthermore, a separate meta-analysis was conducted with studies clustered in subgroups according to different sports performance domains (i.e., endurance, strength, visuomotor skill). Here, studies were divided into these groups depending on the most prominent performance domain in each respective sport studied (e.g., cycling – endurance; bodybuilding – strength; pistol-shooting – visuomotor skill) [12]. According to Cochrane guidelines, pooled standardized mean differences of subgroup analyses and the overall effect were estimated using Cohen’s effect size: small (≤ 0.2), moderate (≤ 0.5) large (≤ 0.8), and very large (> 0.8). The degree of heterogeneity between studies was assessed using Chi^2^ (p < 0.1 considered significant) and the *I*^2^ statistic, with values from ≤ 50% indicating low heterogeneity, 50%–75% moderate heterogeneity, and > 75% high level of heterogeneity, and visual inspection of the funnel plot.

**Table 1.** Overview of studies investigating acute anodal tDCS-induced sport-specific performance changes in trained athletes.

| Study | Design | Participants<br>(F – female, M – male) | Training experience | Anode (A)<br>Cathode (C) | Current<br>intensity (mA) | Current<br>density<br>(mA/cm<sup>2</sup>) | Stimulation<br>duration | Sport-specific<br>motor performance | Outcome |
| --- | --- | --- | --- | --- | --- | --- | --- | --- | --- |
| Okano et al.<br>2015 | Crossover | 10 cyclists (all M) | 10-11 years | A – left TC (T3)<br>C – right supraorbital area<br>(Fp2) | 2 mA | 0.057 | 20 min | Peak power output<br>(PPO) on cycle<br>ergometer | a-tDCS improved<br>PPO compared to<br>sham |
| Holgado et al.<br>2019 | Crossover | 36 cyclists (all M) | Mean VO<sub>2</sub>max = 54<br>mL•min<sup>-1</sup>•kg<sup>-1</sup> | A – PFC (F3)<br>C – right shoulder | 2 mA | 0.08 | 20 min | 20 min TT on cycle<br>ergometer | No significant<br>difference<br>between a-tDCS<br>and sham |
| Kamali, Nami<br>et al. 2019 | Crossover | 8 pistol-shooters (4 F, 4<br>M) | 2- 3 years pistol-<br>shooting | A – right Cerebellum (CB2)<br>C – left dlPFC | 2 mA | 0.057 | 20 min | Shooting score on<br>10 m shooting<br>range | a-tDCS improved<br>shooting score<br>compared to sham |
| Kamali, Saadi<br>et al. 2019 | Crossover | 12 bodybuilders (all M) | 3 training sessions<br>/week | A1 – M1 (Cz, C1, C2)<br>A2 – left TC (T3)<br>C1 – right shoulder<br>C2 – left shoulder | 2 mA | 0.057<br>0.125 | 13 min | Short-term<br>endurance index<br>(SEI): knee<br>extension AMRAP<br>with 30% 1RM | a-tDCS improved<br>SEI compared to<br>sham |
| Mesquita et<br>al. 2019 | Crossover | 19 taekwondo black belts<br>(7 F, 12 M) | 7 taekwondo-specific<br>training sessions,<br>national &<br>international<br>competition | A1 – right M1 (C3)<br>A2 – left M1 (C4)<br>C1 – right shoulder<br>C2 – left shoulder | 1.5 mA | 0.057 | 15 min | Frequency speed of<br>kick test (FSKT) | No significant<br>difference in total<br>number of kicks<br>between a-tDCS<br>and sham |
| Valenzuela et<br>al. 2019 | Crossover | 8 triathletes (all M) | 26-27 hrs/week,<br>International<br>competition | A – left M1 (C3)<br>C – right supraorbital area<br>(Fp2) | 2 mA | 0.08 | 20 min | 800 m freestyle<br>swimming time trial<br>(TT) | No significant<br>difference<br>between a-tDCS<br>and sham |
| Mesquita et<br>al. 2020 | Crossover | 12 taekwondo black belts<br>(4 F, 8 M) | 8.6 years,<br>International &<br>national competition | A1 – right M1 (C3)<br>A2 – left M1 (C4)<br>C1 – right shoulder<br>C2 – left shoulder | 1.5 mA | 0.057 | 15 min | Progressive specific<br>taekwondo test<br>(PSTT) | No significant<br>difference for<br>peak kicking<br>frequency<br>between a-tDCS<br>and sham |
| Pollastri et al.<br>2020 | Crossover | 8 cyclists (all M) | Mean VO<sub>2</sub>max = 72.2<br>mL•min<sup>-1</sup>•kg<sup>-1</sup> | A1 – left dlPFC (F3)<br>A2 – right dlPFC (F4)<br>C1 – Fp1/F7/C3<br>C2 – Fp2/F8/C4<br>(HD-tDCS) | 1.5 mA | n.a. | 20 min | 15 km TT on cycle<br>ergometer | a-HD-tDCS<br>improved TT<br>compared to sham |
| Chen et al.<br>2021 | Crossover | 13 basketball players (all<br>M) | Regular training | A – M1 (Cz)<br>C – M1 (C5, C6)<br>Halo-Sport | 2 mA | 0.083 | 20 min | Power test (counter<br>movement jump) | a-tDCS improved<br>counter movement<br>jump height<br>compared to sham |
| da Silva<br>Machado et<br>al. 2021 | Crossover | 12 endurance athletes (all<br>M, 7 cyclists, 5 rowers) | Mean experience 6.9<br>years, 4.9 hrs/week | A – M1 (Cz)<br>C – inion | 2 mA | 0.056 | 20 min | Time to exhaustion<br>(TTE) test with<br>80% PPO on cycle<br>ergometer | No significant<br>difference in TTE<br>between a-tDCS<br>and sham |
| Fortes et al. 2021 | Crossover | 12 resistance trained athletes (all M) | 3-10 years, 3-5 sessions/week | A – M1 (Cz)<br>C – right shoulder | 2 mA | 0.08 | 20 min | Total number of parallel back squat repetitions with 15 RM over 10 sets | a-tDCS improved total back squat volume compared to sham |
| Grosprêtre et al. 2021 | Crossover | 18 parkour athletes (all M) | Mean training volume: 4343 hrs | A – left M1 (FC2)<br>C – left shoulder<br>A – left dlPFC (F3)<br>C – right supraorbital area | 2 mA | 0.08 | 20 min | Compound jumping performance (standing long jump, squat jump, counter movement jump) | M1-tDCS improved jumping performance compared to sham<br>No significant difference in jumping performance between dlPFC-tDCS and sham |
| Kamali et al. 2021 | Crossover | 14 olympic boxers (all M) | 2 years, 6hrs/week | A1 – right M1 (C3)<br>A2 – left M1 (C4)<br>C1/2 – bilaterally adjacent to spinal processes C5-T1 | 2 mA | 0.125 | 13 min | Boxing specific reaction time task for both hands | a-tDCS improved reaction time of right and left hand compared to sham |
| Penna et al. 2021 | Crossover | 10 swimmers (all M) | 7 hrs/week, competition at highest national level | A – left TC (T3)<br>C – ipsilateral shoulder | 2 mA | 0.057 | 30 min | 800 m swimming TT | No significant difference in the time to complete the 800 m swimming trial between a-tDCS and sham |
| Fortes et al. 2022 | Crossover | 19 swimmers (all F) | 5 years, competition at regional level | A – left oPFC (Fp1)<br>C – right oPFC (Fp2) | 2 mA | 0.08 | 30 min | 3 min all-out tethered swimming | a-tDCS improved force minimum of tethered swimming compared to sham |
| Gallo et al. 2022 | Crossover | 11 cyclists (all M) | Mean $\text{VO}_2\text{max} = 71.8 \text{ mL} \cdot \text{min}^{-1} \cdot \text{kg}^{-1}$ | A1 – left dlPFC (F3)<br>A2 – right dlPFC (F4)<br>C1 – Fp1/F7/C3<br>C2 – Fp2/ F8/C4 (HD-tDCS) | 1.5 mA | n.a. | 20 min | 2 km TT on cycle ergometer | a-tDCS tended to improve 5 km TT compared to sham |
| Liang et al. 2022 | Crossover | 8 rowers (all F) | Competitive performance ranking in top 8 in the national regatta, regular daily training | A – M1 (Cz)<br>C – M1 (C5, C6)<br>Halo-Sport | 2.2 mA | 0.083 | 20 min | 5 km TT on rowing ergometer | No significant difference in 5 km rowing time between a-tDCS and sham |
| Nikooharf Salehi et al. 2022 | Crossover | 15 swimmers (all M) | 6 years, International & national competition | A – left dlPFC (F3)<br>C – right supraorbital area (Fp2) | 2 mA | 0.057 | 20 min | 50 m freestyle swimming trial in mentally fatigued state | a-tDCS improved freestyle swimming performance compared to sham |

## 3 Results

### 3.1 Study selection

The systematic literature search yielded a total of 249 records. After removal of 40 duplicates, 209 records were screened, of which 144 were excluded based on title and abstract. The remaining 65 records were assessed for eligibility. Based on PICOS criteria, 48 records were excluded due to the following reasons: no homogeneous athlete sample (n = 24), no RCT (n = 9), no sport-specific motor task was performed (n = 13), article not in English (n = 1), no single tDCS-Session (n = 1). In addition, a Google Scholar search identified one study that was not indexed in the searched databases because of its recent publication date. Finally, a total of 18 studies were deemed eligible for inclusion in the qualitative synthesis [3, 16, 17, 29-43]. An overview of the study selection process is depicted in the PRISMA flow diagram (Figure 1).

### 3.2 Risk of bias assessment

The risk of bias was found to be low in most of the studies reviewed. However, 8 of the 18 included studies (44%) were single-blind trials, which poses a risk of detection bias due to problems with blinding of the outcome assessment. A summary of the risk of bias assessment is visualized in Figure 2.

**Figure 2.**
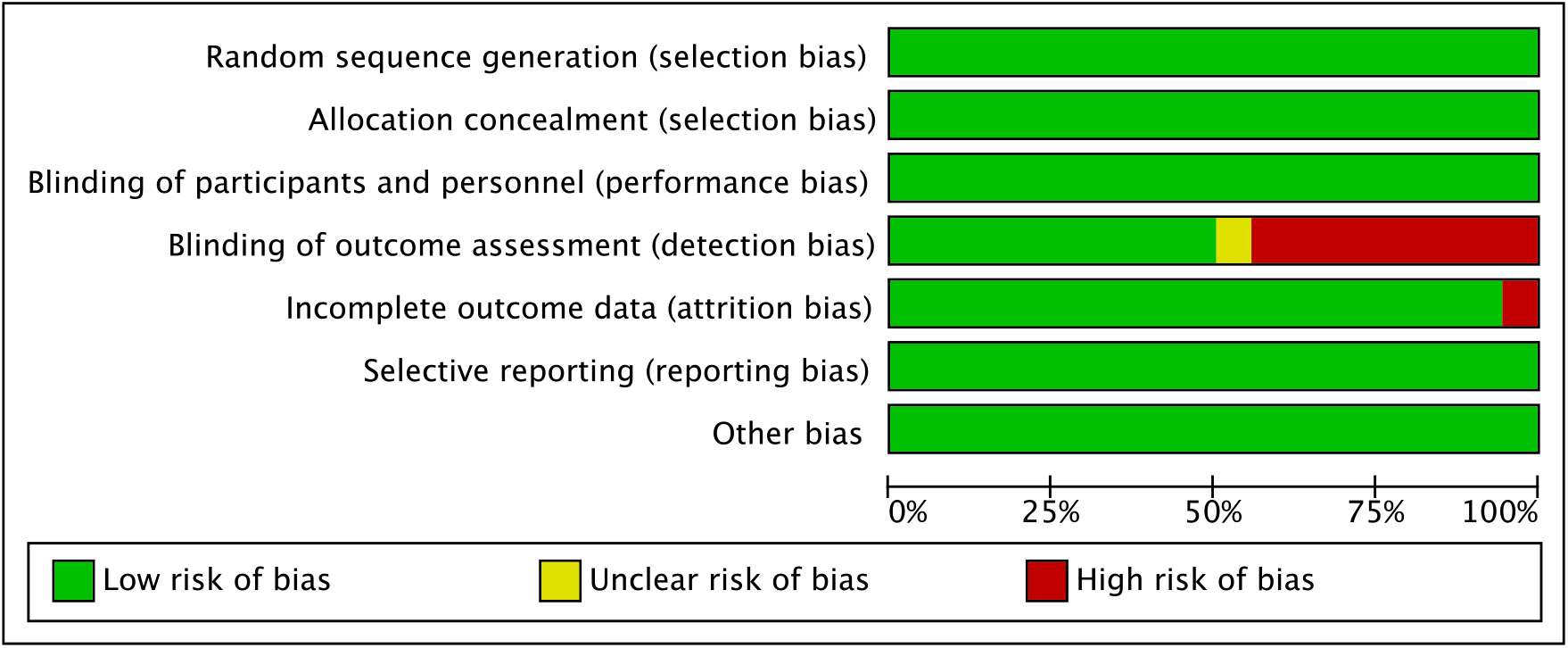
Risk of bias assessment according to Cochrane guidelines. The checklist consists of 7 items, with the following answers choices: “low risk of bias” (“+”), “high risk of bias” (“-”), or “unclear risk of bias” (“?”).

**Figure 3.**
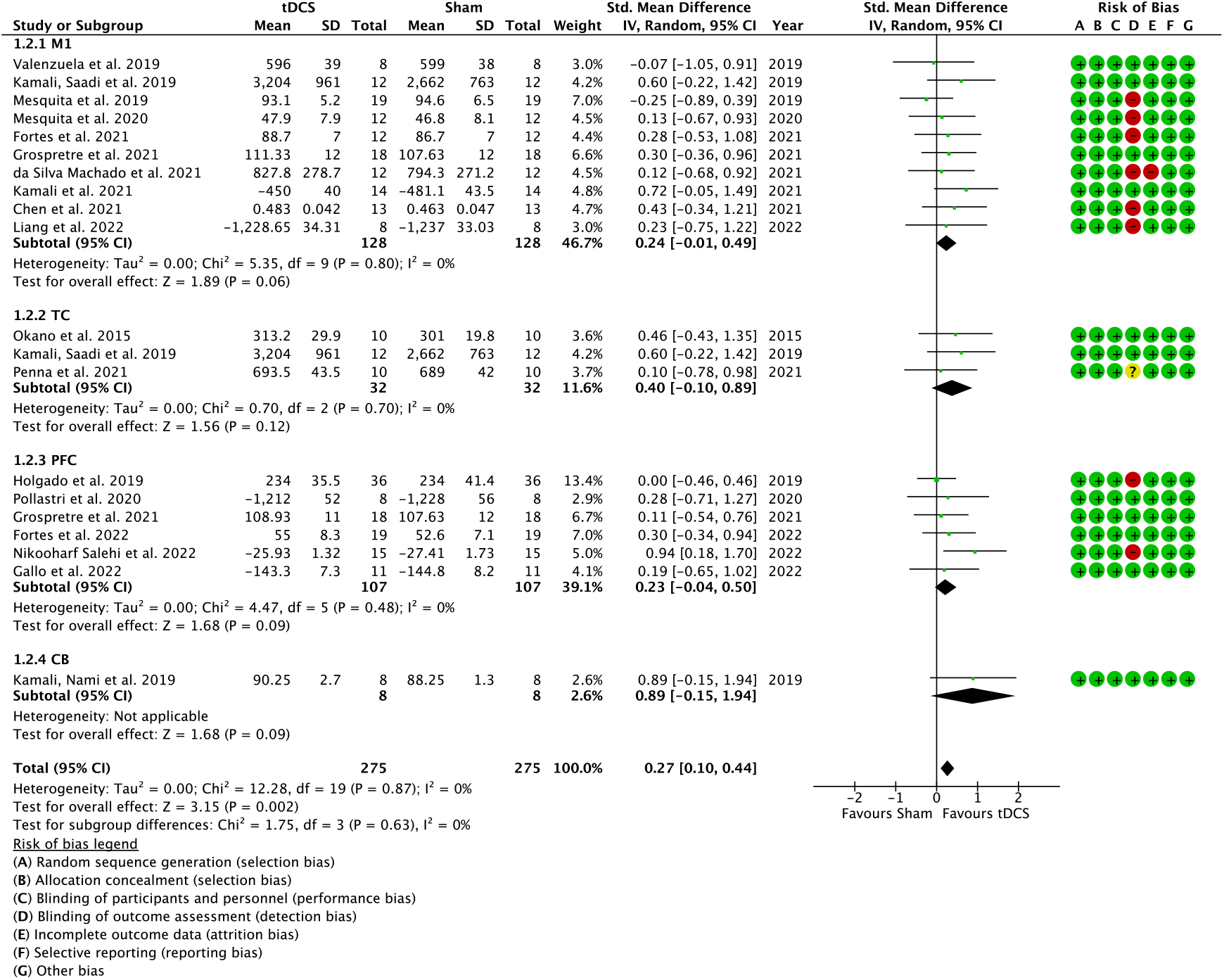
Forest plot showing standardized mean difference (SMD) for comparing sport-specific performance changes in trained individuals between anodal tDCS and sham. Studies were clustered in subgroups according to the cortical target area of tDCS, i.e., motor cortex (M1), temporal cortex (TC), prefrontal cortex (PFC), and cerebellum (CB).

**Figure 4.**
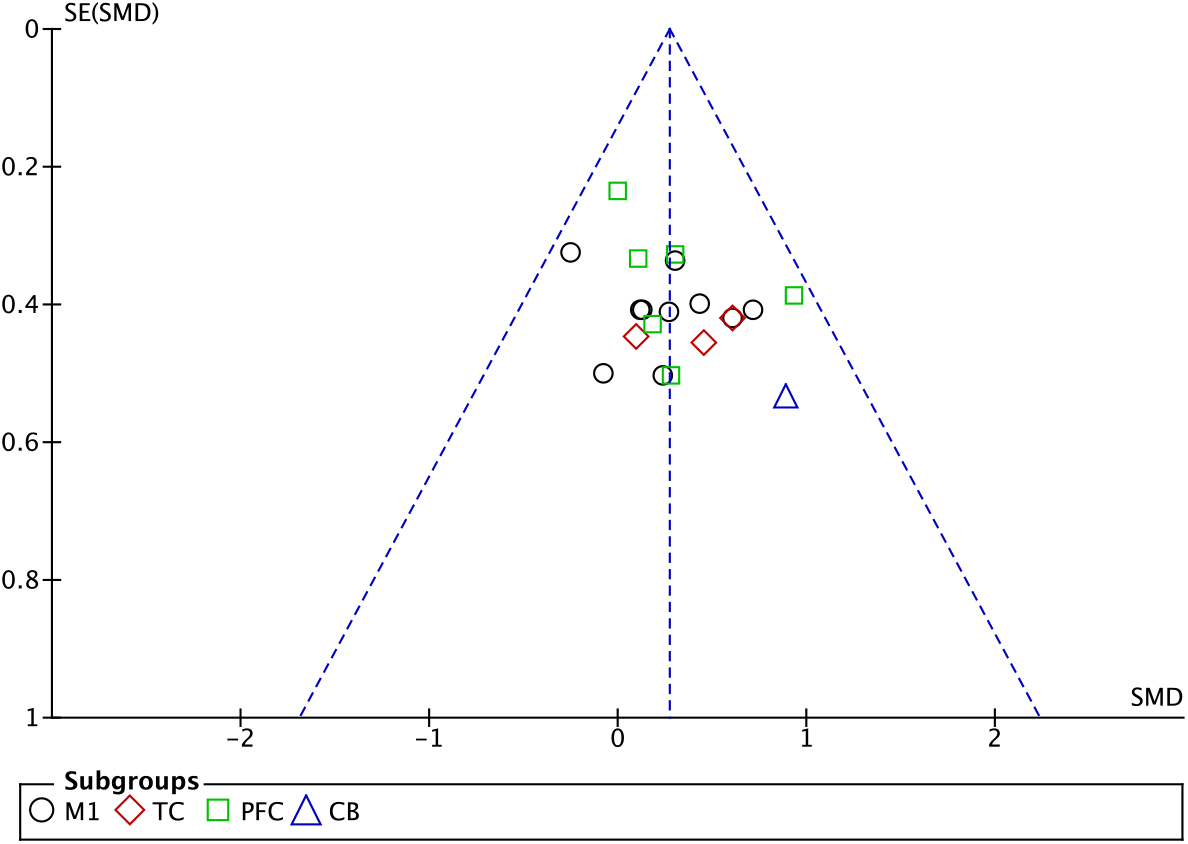
Funnel plot of studies included in the quantitative meta-analysis divided into subgroups depending on the brain target of tDCS. Effect sizes are scattered symmetrically and lie within the funnel.

**Figure 5.**
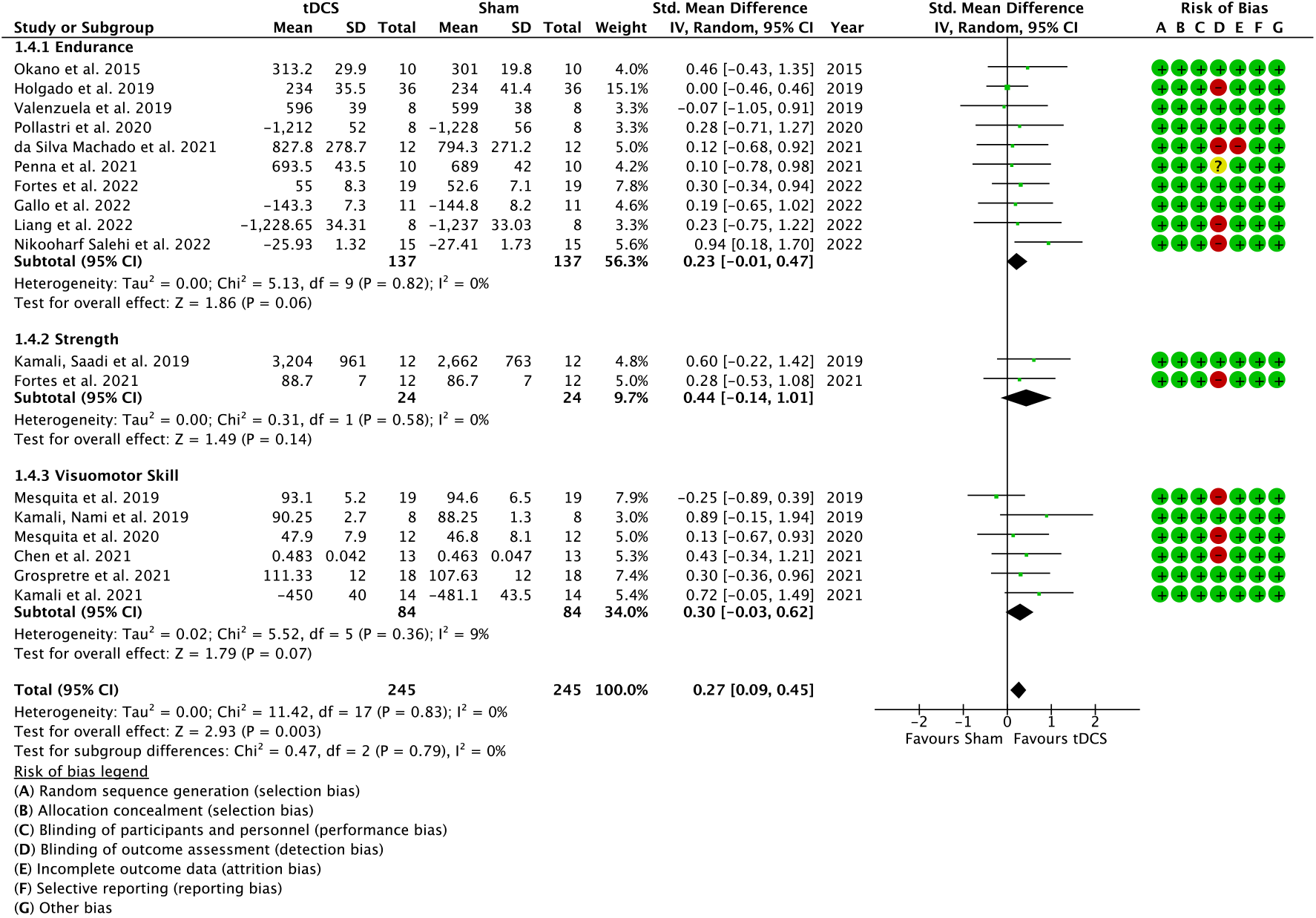
Forest plot showing standardized mean difference (SMD) for comparing sport-specific performance changes in trained individuals between anodal tDCS and sham. Studies were clustered in subgroups depending on the most prominent performance indicator in each respective sport studied (e.g., cycling – endurance; bodybuilding – strength; pistol-shooting – visuomotor skill).

### 3.3 Study design and participant characteristics

A comprehensive summary of study characteristics is presented in Table 1. All included studies were designed as crossover RCT with sham as a comparator and published between 2015 and 2022. Interestingly, the majority of studies (67 %) were published in the last 2 years, highlighting the recent interest in this area of research. In total, 245 trained athletes (mean age range: 19 – 33 years) were enrolled in these studies, with 203 being male and 42 being female. Sample sizes of included studies ranged from 8 to 36 participants (13.6 ± 6.6). Athletes were experienced in various sports disciplines: cycling [16, 29, 35, 36, 41], swimming [37, 39, 43], triathlon [30], rowing [41, 42], bodybuilding [31], resistance training [38], basketball [40], parkour [3], taekwondo [17, 32] and pistol-shooting [33]. Additionally, some studies reported that athletes competed at the regional [39], national [17, 32, 37, 42, 43] and/or international level [17, 30, 32, 37]. For an overview of investigated sport-specific motor performances in each study, please see Table 1. Sport disciplines were categorized in the following performance domains: endurance (cycling [16, 29, 35, 36, 41], swimming [37, 39, 43], triathlon [30], rowing [41, 42]), strength (bodybuilding [31], resistance training [38]) & visuomotor skill (basketball [40], parkour [3], taekwondo [17, 32], pistol-shooting [33]).

Only anodal tDCS conditions from experiments in which stimulations with multiple polarities were applied were analyzed in the current systematic review and meta-analysis due to our study aims of investigating performance-enhancing effects. For tDCS application, the following electrode montages were used: conventional tDCS (one pair of sponge electrodes), bilateral conventional tDCS (two pairs of sponge electrodes), Halo-Sport device (headphones with two implemented electrodes), and HD-tDCS (arrays of small gel-based electrodes). Conventional tDCS was used in 10 studies (56 %), bilateral conventional tDCS in 4 studies (22 %), the Halo-Sport device in 2 studies (11 %) and HD-tDCS in 2 studies (11 %). One study compared conventional tDCS with HD-tDCS [41]. Because there were no differences between conventional tDCS and HD-tDCS in this study, the focus of the current review was on conventional tDCS. Additionally, a single study combined conventional tDCS with transcutaneous spinal current stimulation [34]. All studies applied tDCS prior to sport-specific performance. Cortical target areas of tDCS were clustered in 4 groups: M1 [3, 17, 30-32, 34, 38, 40-42], TC [29, 31, 43], PFC [3, 16, 35-37, 39] and CB [33]. Current intensity was set at 2 - 2.2 mA in most studies (78 %) or 1.5 mA (22%). Current was applied for a duration of either 20 min (67 %), 12-15 min (22 %), or 30 min (11 %).

### 3.4 Quantitative Analysis

#### 3.4.1 Overall Effect

Across all included studies, a significant moderate standardized mean difference favoring anodal tDCS over sham for sport-specific motor performance changes could be observed (SMD = 0.27, 95%CI [0.10, 0.44], p = 0.002). Studies showed low heterogeneity (Chi^2^ = 12.28, p = 0.87; I^2^ = 0%). In addition, the funnel plot showed symmetrically scattered effect sizes, all within the funnel.

#### 3.4.2 Cortical target area subgroup analysis

Subgroup analysis depending on cortical target areas of tDCS indicated non-significant moderate to high standardized mean differences in favor of anodal tDCS compared to sham for M1 (SMD = 0.24, 95%CI [-0.01, 0.49], p = 0.06, Chi^2^ = 5.35, p = 0.80; I^2^ = 0%), TC (SMD = 0.40, 95%CI [-0.10, 0.89], p = 0.12, Chi^2^ = 0.70, p = 0.70; I^2^ = 0%), PFC (SMD = 0.23, 95%CI [-0.04, 0.50], p = 0.09, Chi^2^ = 4.47, p = 0.48; I^2^ = 0%) and CB (SMD = 0.89, 95%CI [-0.15, 1.94], p = 0.09, n.a.). No between subgroup differences were observed (Chi^2^ = 1.75, df = 3, p = 0.63).

#### 3.4.3 Performance domain subgroup analysis

Performance domain subgroup analysis revealed non-significant moderate standardized mean differences favoring anodal tDCS compared to sham for sport-specific performance changes: endurance domain (SMD = 0.23, 95%CI [-0.01, 0.47], p = 0.06, Chi^2^ = 5.13, p = 0.82; I^2^ = 0%), strength domain (SMD = 0.44, 95%CI [-0.14, 1.01], p = 0.14, Chi^2^ = 0.31, p = 0.58; I^2^ = 0%) and visuomotor skill domain (SMD = 0.30, 95%CI [-0.03, 0.62], p = 0.07, Chi^2^ = 5.52, p = 0.39; I^2^ = 9%). Also, no between subgroup differences were observed (Chi^2^ = 0.47, df = 2, p = 0.79).

## 4 Discussion

We investigated the effects of tDCS on sport-specific performance changes in athletes. The idea of this study relates to the fundamental debate regarding the scope of applicability of tDCS, in other words, the ecological validity of tDCS. Concerning motor performance, tDCS is predominantly employed in patients or healthy participants with proven ability to enhance motor performance [5-8]. However, the question remains whether performance-enhancing effects are also detectable in trained athletes, particularly in sport-specific motor tasks. Due to so-called “ceiling effects”, it is assumed that performance enhancement becomes increasingly difficult to achieve as the level of performance increases [41]. A total of 18 studies were included in this meta-analysis. Overall, our results show a moderate effect of anodal tDCS on sport-specific performance changes in athletes compared to sham. To provide a better classification of tDCS effects, two additional subgroup analyses were performed, one on cortical target areas and one concerning the performance domain. Neither subgroup analysis revealed a significant effect. All results and their implications are discussed below.

To contextualize the observed effect of tDCS on sport-specific performance in athletes, the variability of the respective studies must be taken into account. In principle, there is no consensus concerning the mode of action of tDCS as a function of specific stimulation sites. However, certain areas of the brain are known to play a role in the execution and control of athletic performance such as the prefrontal cortex (PFC), primary motor cortex (M1), temporal cortex (TC), and the cerebellum (CB). In healthy populations, anodal tDCS over M1 has been shown to enhance muscle strength in some studies [15, 44, 45], while others fail to replicate such effects [46, 47]. Similarly, mixed results exist concerning tDCS effects on endurance [48-50]. Our results corroborate this heterogeneity for sport-specific performance in athletes. Among the ten included studies that stimulated M1, five showed performance-enhancing effects [3, 31, 38, 40], whereas the remaining five studies failed to demonstrate any behavioral effects [17, 30, 32, 41, 42]. On a functional level, anodal tDCS over M1 has been shown to increase the excitability of M1 [4], which improves the neural drive to the working muscles [51]. An upregulation of neural output can lead to improvements in physical performance, especially in the strength domain, through enhanced utilization of neuromuscular capacities. This mechanism may constitute a major factor driving the observed increases in strength measures as a result of anodal tDCS over M1 [31, 38]. Furthermore, a prolongation of the onset of so-called central fatigue by M1 stimulation is also conceivable [52]. Reduced or ceased firing of motor units contributes to the loss of muscle function associated with central fatigue [53]. This process can be counteracted by tDCS via a delay of the motor slowing effect, i.e., a reduction in movement speed during fast repetitive movements [21]. A similar effect was observed in one of the studies included in our meta-analysis. [40] examined the repeated-sprint ability and observed an improvement following anodal tDCS over M1. Another aspect of performance that is potentially mediated through M1 stimulation is pain tolerance. Previous research demonstrated that M1 stimulation can increase pain perception thresholds in healthy individuals [54]. It has been suggested that individuals with better pain tolerance are more successful in their sport [55], which is related to fatigue, as fatigue is facilitated by lower corticospinal excitability as well as decreasing pain tolerance [30]. One study of elite athletes showed that, compared to non-athletes, elite athletes had higher pain tolerance, higher heat pain thresholds, and lower perceived pain intensity with thermal stimulation [56]. Accordingly, M1 stimulation may also enhance athletic performance due to an attenuation of exercise-induced pain.

Athletic performance necessitates the adaptation of autonomic physiology to external demands. The central autonomic network ensures such adaptations [57]. This network governs the autonomic nervous system through the integration of higher cortical centers to adapt the system to specific demands (cortical component) [58], while also comprising sympathetic and parasympathetic sections within the brainstem involved in monitoring the physiological status quo via baro-and chemoreceptor afferents (subcortical component) [57]. Essential parts of this network include the temporal cortex (TC) and the insular cortex (IC). For instance, the TC represents a higher-order control of cardiac autonomic functions [59], whereas the IC acts as a central interface between cortical and subcortical components of the central autonomic network [57]. Two studies included in our analysis stimulated TC [29, 43], while another study used a dual stimulation setup of M1 and TC [31]. In a seminal study, [29] demonstrated a positive effect of anodal tDCS over TC on performance in a maximal incremental exercise test on a bicycle ergometer. Notably, the authors showed that the performance enhancement was due to a delay in vagal withdrawal, suggesting a potential link between TC and control of autonomic cardiac functions, and also their susceptibility to alteration by tDCS. Following the results of [29], [31] also found positive tDCS-induced effects on endurance and strength performance of bodybuilders following dual-stimulation of M1 and TC. Again, these results were associated with vagal withdrawal, confirming the previous findings. Another study failed to observe differences in swimming performance following TC stimulation [43]. However, autonomic cardiac functions were not monitored, which complicates potential explanations concerning the absence of an effect. For this purpose, future studies aiming to stimulate TC should always monitor autonomic cardiac functions to be able to draw conclusions on the origin of potential performance enhancements.

Another area involved in exercise regulation is the prefrontal cortex (PFC). Functional roles of prefrontal subdivisions such as dorsolateral prefrontal cortex (dlPFC) and orbital prefrontal cortex (oPFC) extend from cognitive control of motor behavior [60] to the disengagement of motor activity [61], and fatigue [62]. Six studies included in this meta-analysis stimulated the PFC [3, 16, 35-37, 39]. Given the inhibitory control of the PFC during motor activity, a common rationale of studies aiming to employ PFC stimulation is based on the assumption that an upregulation of PFC excitability leads to a reduction in effort for inhibitory control during motor activity. In this sense, the perceived effort during exercise would be reduced and the termination of exercise would be postponed [36]. Evidence for this can be found in studies demonstrating that sensory signals relating to the perception of effort are processed by areas functionally associated with the PFC, such as the supplementary motor area (SMA), premotor cortex (PMC), and M1 [63]. None of the included studies that examined perceived exertion found any modulatory effects between PFC stimulation and sham [16, 35, 36]. However, in all three studies, motor performance also increased. Hence, this finding might indicate an improved inhibitory control during exercise after anodal PFC stimulation, as inhibitory control moderated by prefrontal areas may contribute to the overall perception of effort during exercise [63]. tDCS may have reduced the cognitive effort needed to exert inhibitory control, allowing for higher levels of performance with the same perceived effort [36]. Another notable aspect of PFC functioning is the fact, that the ability to maintain PFC oxygenation at high exercise intensity is related to better endurance performance [64]. Moreover, PFC oxygenation decreases before the onset of fatigue [65], highlighting the importance of the PFC in the cognitive regulation of motor activity. With the exception of one study [16], all other studies that stimulated PFC showed an increase in motor performance in the endurance domain. Although no definitive conclusions can be drawn, it is, therefore, tempting to speculate that anodal stimulation of the PFC may delay the termination of motor activity by increasing the ability of the PFC to temporarily disregard effort-related cues and maintain a constant neural motor drive.

Finally, another brain region that plays an essential role in motor control is the CB. One aspect of cerebellar motor control relates to the so-called forward model [66]. Specifically, this model outlines the idea that the cerebellum receives a copy of the motor command and computes the sensory consequences of that command through input from the periphery [67]. Thus, the model provides a solution for dynamic adaptation of motor commands based on sensory consequences [68]. It follows that cerebellar tDCS is predominantly employed with the goal of reducing errors during motor tasks. Previous studies observed a reduction in movement errors in various tasks, with improvements mainly attributed to postural adjustments resulting from cerebellar tDCS [69, 70]. In the sole study included within the present meta-analysis, the shooting accuracy of pistol shooters was increased by anodal stimulation of the cerebellum [33]. The authors attributed this to rapid postural adaptations that allowed the shooters to reduce physiological tremor. Since postural adjustments are a necessary foundation for athletic performance, future studies should examine the efficacy of cerebellar tDCS on sports-specific performance. The limited number of studies on cerebellar tDCS and performance enhancement highlights the potential to examine the efficacy of cerebellar tDCS in this area in the future.

In summary, the following observations may be noted. Of 18 included studies, 10 studies investigated effects in the endurance domain. Five of these studies found an increase in specific endurance performance (PFC (n=4); TC (n=1)) while five could not demonstrate such effects (PFC (n=1); M1 (n=3); TC (n=1)). Two studies examined anodal tDCS effects on sport-specific strength performance and demonstrated improved performance in training volume at fixed load levels (M1 (n=2)). The remaining 6 studies examined potential increases in specific visuomotor skills. Four studies were able to demonstrate positive anodal tDCS effects (M1 (n=3); CB (n=1)) whereas two studies did not observe such effects (M1 (n=2)). Although some trends can be observed (e.g., the tendency that anodal stimulation of the PFC, with the exception of one study, leads to an increase in sport-specific endurance performance in athletes), no significant insights into the specificity of tDCS in the context of sport-specific performance enhancement in athletes can be stated.

### Limitations

Compared to sham stimulation, anodal tDCS administered over specific brain areas can induce performance-enhancing effects. This has been demonstrated regularly in non-athletes, recreationally active individuals, and even in high-level athletes. However, we did not observe any differences in the effectiveness of tDCS, neither concerning the site of stimulation nor between different physical domains. These findings may relate to the known variability problem of tDCS applications, meaning participants either show or do not show improvements in certain behavioral parameters [71]. Origins for this phenomenon range from differences in age, gender, anthropometry, and genetics, to morphological differences, cortical excitability, attention, time of day, as well as stimulation parameters, such as current density and electrode size [72, 73]. In addition, the lack of subgroup effects can also be attributed to insufficient statistical power. Since relatively few studies have investigated tDCS effects in athletes focusing on sport-specific performance enhancements, future studies should focus on this issue more thoroughly. It remains to be seen whether the trend of using tDCS for performance enhancement in competitive sports will continue in the coming years. A final limitation concerns the classification of the performance domains. Here, our classification was an initial attempt to categorize tDCS-related effects on athletic performance to delineate the range of tDCS effects. However, the unambiguous assignment of each sport to one of these categories is unrealistic. For this reason, we decided to use not the sport but the sport-specific motor task employed in the included studies as the starting point of our categorization. In the future, it seems reasonable to design tDCS studies with realistic sport-specific conditions in mind. This will allow for a better classification of potential performance-enhancing effects and a more precise understanding of the mechanisms of tDCS in the context of sports performance.

## 5 Conclusions

In conclusion, a single anodal tDCS session on cortical areas relevant to motor function leads to performance enhancement of athletes in sport-specific tasks. Although no conclusions can be drawn regarding the modes of action as a function of performance domain or stimulation site, these results imply intriguing possibilities concerning sports performance enhancement. A fundamental novelty of our approach is the concept that performance enhancement in high-level athletes must also be studied in sport-specific, naturalistic settings. Apart from ethical considerations, our results can be considered as a starting point for future research on the performance enhancement of athletes by tDCS. It remains to be seen what trend future results will reveal, but the potential of this method for sports performance enhancement does not seem to be exhausted.

## Data Availability

All data produced in the present work are contained in the manuscript

